# Protection of homologous and heterologous boosters after primary schemes of rAd26-rAd5, ChAdOx1 nCoV-19 and BBIBP-CorV during the Omicron outbreak in adults of 50 years and older in Argentina: a test-negative case-control study

**DOI:** 10.1101/2022.09.25.22280341

**Authors:** Soledad González, Santiago Olszevicki, Alejandra Gaiano, Martín Salazar, Lorena Regairaz, Ana Nina Varela Baino, Erika Barkel, Teresa Varela, Veronica V. González Martínez, Santiago Pesci, Lupe Marín, Juan Ignacio Irassar, Leticia Ceriani, Enio Garcia, Nicolás Kreplak, Elisa Estenssoro, Franco Marsico

## Abstract

**Objectives:** To estimate the protection against laboratory-confirmed SARS-CoV-2 infection, hospitalisations, and death after homologous or heterologous third-dose (booster) in individuals with primary vaccination schemes with rAd26-rAd5, ChAdOx1nCoV-19, BBIBP-CorV or heterologous combinations, during the period of Omicron BA.1 predominance.

**Design:** Retrospective, test-negative, case-control study, with matched analysis.

**Setting:** Province of Buenos Aires, Argentina, between 12/1/21-4/1/21.

**Participants:** 422,144 individuals ≥50 years who had received two or three doses of COVID-19 vaccines and were tested for SARS-CoV-2.

Main outcome measures: Odds ratios of confirmed SARS-CoV-2 infection, hospitalisations and death after administering different boosters, compared to a two-dose primary scheme.

**Results:** Of 221,933(52.5%) individuals with a positive test, 190,884(45.2%) had received a two-dose vaccination scheme and 231,260(54.8%) a three-dose scheme. The matched analysis included 127,014 cases and 180,714 controls.

The three-dose scheme reduced infections (OR 0.81[0.80-0.83]) but after 60 days protection dropped (OR 1.04[1.01-1.06]). The booster dose decreased the risk of hospitalisations and deaths after 15-59 days (ORs 0.28[0.25-0.32] and 0.25[0.22-0.28] respectively), which persisted after administration for 75[66-89] days.

Administration of a homologous booster after a primary scheme with vectored-vaccines provided low protection against infections (OR 0.94[0.92-0.97] and 1.05[1.01-1.09] before and after 60 days). Protection against hospitalisations and death was significant (OR 0.30[0.26-0.35] and 0.29[0.25-0.33] respectively) but decreased after 60 days (OR 0.59[0.47-0.74] and 0.51[0.41- 0.64] respectively).

The inoculation of a heterologous booster after a primary course with ChAdOx1 nCoV-19, rAd26-rAd5, BBIBP-CorV, or heterologous schemes, offered some protection against infection (OR 0.70[0.68-0.71]), which decreased after 60 days (OR 1.01[0.98-1.04]). The protective effect against hospitalisations and deaths (OR 0.26[0.22-0.31] and 0.22[0.18-0.25] respectively) was clear and persisted after 60 days (OR 0.43[0.35-0.53] and 0.33[0.26-0.41]).

**Conclusions:** This study shows that, during Omicron predominance, heterologous boosters provide an enhanced protection and longer effect duration against COVID-19-related hospitalisations and death in individuals older than 50, compared to homologous boosters.

## Introduction

The emergence of the highly transmissible omicron (B.1.1.529) variant of concern (VOC), able to partially evade the immune response achieved after vaccination or natural infection, has caused an extraordinary increase in COVID-19 cases worldwide.^1^ As evidence of waning of the immunity generated by mRNA vaccines began to surface, many countries started to administer a booster to improve vaccine response against omicron at the end of 2021.^2^ Vaccine Effectiveness (VE) can be restored with a booster dose. Thus far, most reports about boosters refer to the administration of the same mRNA vaccines administered as primary schemes.^3,4^

Argentina started the massive vaccination roll-out on December 29, 2020, with the recombinant adenovirus (rAd)-based vaccine rAd26-rAd5 (Sputnik V, from Gamaleya National Research Center for Epidemiology and Microbiology). In a context of decreased vaccine availability across the world, the Argentine Ministry of Health incorporated other immunisation schedules, which included: the vectored vaccines ChAdOx1 nCoV-19 (from Oxford University and AstraZeneca) and CanSinoBIO Ad5-nCoV-S (from CanSino Biologics Inc), the inactivated viral SARS-CoV-2 vaccine BBIBP-CorV (from Beijing Institute of Biological Products Co); and the mRNA vaccines BNT162b2 (from Pfizer-BioNTech) and mRNA-1273 (from Moderna). To achieve wide vaccine coverage in the shortest possible time, Argentina started the use of heterologous vaccination schemes in July 2021. Recommendations about this strategy are available in the literature; furthermore, the advantages of applying heterologous boosters to improve the immunological response against variants of concern (VOCs), including omicron, has been demonstrated in experimental and real-world studies.^4–21^

There is scarce information in real-world studies about the protection achieved and duration of homologous or heterologous boosters following a primary vaccination course with the BBIBP-CorV, rAd26-rAd5 or with heterologous vaccines.^9,21^ Therefore, our aim was to estimate the protection against laboratory-confirmed SARS-CoV-2 infection, hospitalisations, and death after homologous or heterologous booster doses in individuals that had previously received rAd26-rAd5, ChAdOx1 nCoV-19, BBIBP-CorV or heterologous schemes as primary series vaccination during a period of omicron BA.1 predominance.

## Methods

### Study population and design

This study used a test negative case-control design, which has proven to limit bias resulting from testing and healthcare seeking behaviour.^22,23^ Subjects eligible for inclusion were ≥50 years with residence in the Province of Buenos Aires, had received at least two doses of COVID-19 vaccines by 1 January 2022, and were tested for SARS-CoV-2 between 1 January – 1 April, 2022. Exclusion criteria were having a previous positive RT-PCR or antigen tests for SARS-CoV2 in the previous 90 days, having received none, one, or four doses of any vaccine, or having a laboratory-confirmed test occurring within 14 days of vaccination.

We assessed vaccine performance during the period of omicron B.1.1.529 predominance (1 January-1 April 2022), as detected by the National Ministry of Health’s genomic surveillance program for identifying VOCs through RT-PCR laboratory-confirmation.^24^

### Data sources and definitions

This study used epidemiological surveillance data from the National Surveillance System (SNVS 2.0). The database registered age, gender, presence or absence of comorbidities and site of residence (Greater Buenos Aires or not). Information on SARS-CoV-2 infections was obtained using RT-PCR or antigen test until 1 April 2022. During the study period only symptomatic cases were tested, according to the standards established by the Province of Buenos Aires. Information about hospitalisations and death was recorded until 28 April 2022.

The date of confirmed-laboratory SARS-CoV-2 infections was identified by symptom-onset date or, if not available, the date of the sample collected for the COVID-19 test. Number of positive tests in the past, total number of previous tests, dates of hospitalisations and death were registered.

The first positive test during the study period was considered a case for primary analysis, regardless of the number of previous negative tests. Controls were those individuals who tested negative over the entire study period, and were selected according to the date of their first test. Individuals could be included only once for each outcome.

Vaccination information was collected in VacunatePBA, a system developed in the Province of Buenos Aires, Argentina, to address the rollout of the COVID-19 vaccination campaign. It included vaccination date, number of doses, vaccine type, vaccine lot number, and vaccination center. Vaccination status was verified on the day the SARS-CoV-2 test was performed.

In December 2020, Argentina started the vaccination campaign against COVID-19 with rAd26-rAd5 and, progressively incorporating ChAdOx1 nCoV-19, BBIBP-CorV, Ad5-nCoV, BNT162b2, mRNA-1273 vaccines, and combinations in settings of limited vaccine availability rAd26/mRNA1273, rAd26-/ChAdOx1 nCoV-19, and rAd26/Ad5-nCoV among others.^25,26^

The primary vaccination series initially consisted of two doses with a minimum interval of 21- or 28-day for immunocompetent subjects, or three doses with a 28-day interval for immunocompromised adults.^25^ Due to low availability of vaccines, in March 2021 the first dose of viral-vectored vaccines (rAd26-rAd5 and ChAdOx1 nCoV-19) was prioritised, which consequently delayed the second dose for at least 90 days. The interval between doses with the BBIBP-CorV vaccine was left at 28 days.^27^

On 28 October 2021, the WHO recommended an additional dose for individuals aged >50 who had received a primary series with inactivated vaccines.^28^ On 10 November, a booster dose with either rAd26, ChAdOx1 nCoV-19, Ad5-nCoV, BNT162b2 or a half dose (50 μg) of mRNA-1273 was introduced for adults >70 and for those in high-risk groups.^29^

The recommended interval between the initial scheme and the booster was 6 months; afterwards, with the emergence of new evidence, it was shortened to 4 months.^30^ Following the comorbidity-prioritised and age-progressive COVID-19 vaccination campaign, the program continued in a staggered manner, descending in 10-year age decrements, until covering the entire population.

In this study, only individuals >50 were considered, as they were prioritised for vaccination in the guidelines proposed by the National Ministry of Health due to the increased risk of severe disease and mortality demonstrated in this age group.^31,32^ For study purposes, eligible individuals were those who had received a two-dose vaccination scheme and should have received their booster dose equal or over 120 days, but had not—for any reason. They constituted the reference group. Individuals ineligible for boosters were those who had received a two-dose scheme with the last dose administered up to 119 days before the test.

The booster was considered homologous when the platform was similar to that of the primary scheme administered, and heterologous when the platform was different. The analysis of time since vaccination was stratified in two periods of ≤ 60 and ≥ 60 days, taking into consideration the reported increase in COVID-19 vaccine protection after administration followed by a waning over time.^14,18^

The COVID-19 vaccine uptake in the Province of Buenos Aires adults aged 50 and older and the epidemiological characteristics are shown in the Supplementary Material, figure S1,S2.

### Outcomes

The main outcome was the odds ratio of SARS-CoV-2 infection, hospitalisations and death of administering a booster in comparison to a two-dose primary scheme, occurring ≥14 days after the booster dose. The secondary outcome was the odds of experiencing infection, hospitalisations and death related to the administration of homologous and heterologous boosters administered after different primary vaccination schemes.

### Statistical analyses

Data are expressed in tables as mean ± standard deviations, median with 0.25 and 0.75 percentiles or numbers and percentages, as appropriate. T tests, Chi-square tests, Kruskal-Wallis and Wilcoxon-Mann-Whitney tests were used, according to the nature of the variables. A p value<0.05 was considered significant.

The matching process for the test negative case design was performed without replacement using the nearest neighbour (1nn) matching methodology, by means of a logistic regression propensity score within groups defined by exact coincidence on the number of positive tests in the past, gender, site of residence (Greater Buenos Aires or not), and presence or absence of comorbidities. Additionally, we matched the total number of previous tests on three levels (0, 1-2 or 3+) as a proxy of differences in exposure. The non-exact variables considered were age at diagnosis and date of testing, with maximum tolerance of ±2 years for the age and ±6 days for the date of testing. Up to five controls per case were selected.

Subsequently, for each matched set, we utilised a conditional univariate logistic regression model to estimate the odds ratio for the outcome for each of the groups in the vaccination status variable. This variable had four levels. The reference group were those individuals with two doses who were eligible for receiving a booster dose. The other groups included individuals with two doses ineligible for booster, as defined previously; those with three doses, the last received 15-59 days before the test and those with three doses, the last received 60 or more days before the test.

Data preprocessing was carried out with PostgreSQL (Portions Copyright © 1996-2022, The PostgreSQL Global Development Group). All statistical analyses were performed with R (R Development Core Team, 4.2.1 version) software.

## Data Availability

All data produced in the present study are available upon reasonable request to the authors

## Ethics

The Central Ethics Committee of the Ministry of Health of the Province of Buenos Aires evaluated and approved the protocol of the present study on 21 September 2022. The report number is 2022-31701807-GDEBA-CECMSALGP. Informed Consent.This study was exempted of informed consent due to its retrospective nature, and given it is a public health-related official program. Anonymisation of data. Data were anonymized by the following procedure: the personal ID number was used to link the databases of follow-up and of vaccination. After this process, the personal ID number was removed and an ID reference number for each individual was created. This reference number is not associated with any personal information.

## Patient and public involvement

No patients or members of the public were directly involved in the development or completion of this study owing to time and funding constraints.

## Results

### Description of the study population

During the study period, 422,144 subjects aged ≥50 were eligible for the study and obtained one test for SARS-CoV-2 at least once during the period of 1 January to 1 April 2022. Of them, 221,933 (52.5%) individuals had a positive test and 200,211 (47.5%) obtained a negative test. With respect to their vaccination status, 190,884 (45.2%) had received a two-dose scheme of SARS-CoV-2 vaccines and 231,260 (54.8%) a primary scheme plus a booster dose (three-dose scheme). The flowchart of the study is shown in figure 1.

**Figure 1:**
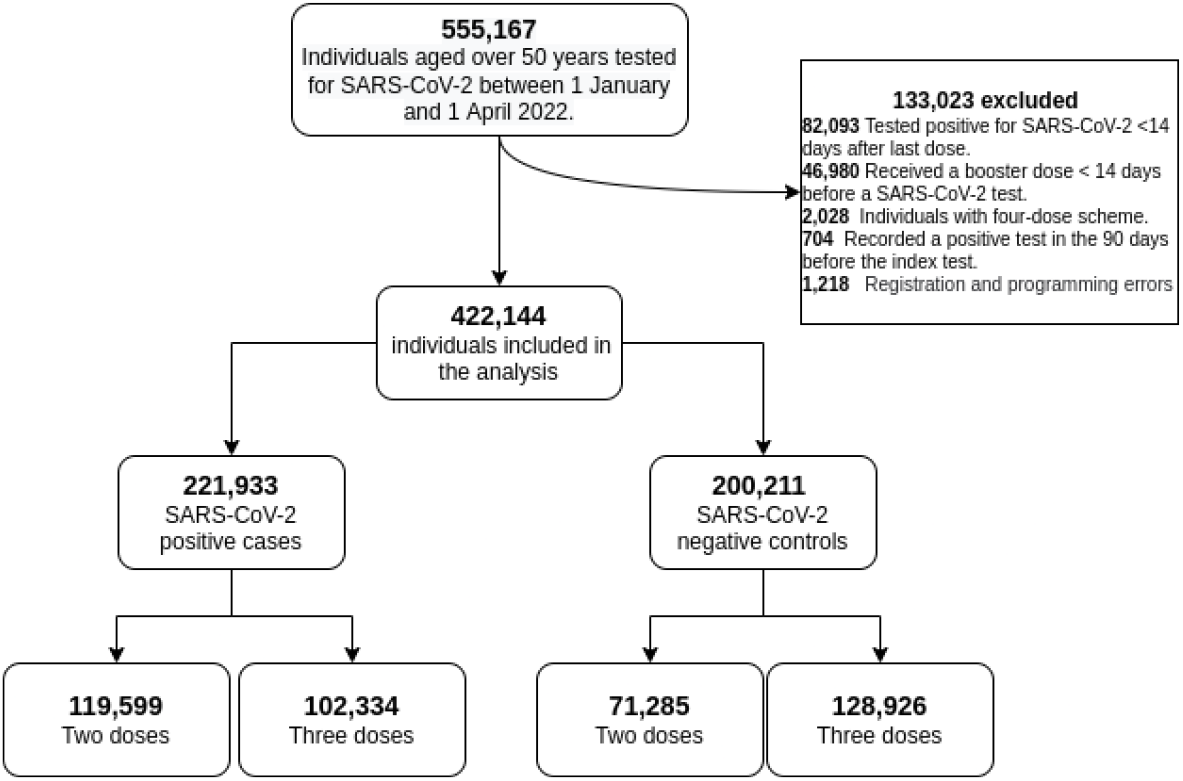
Flowchart of the study

The primary series most frequently included were: ChAdOx1 nCoV-19/ChAdOx1 nCoV-19 (n=143,721; 34.0%), rAd26-rAd5 (n=119,441; 28.3%), BBIBP-CorV/BBIBP-CorV (n=68,251; 16.2%), rAd26/mRNA1273 (66,157; 15.7%) and rAd26-/ChAdOx1 nCoV-19 (n=23,098; 5.5%). The boosters applied were vectored vaccines (n=161,619, 38.3%), mRNA (n=68,850, 16.3%), and other types of booster platforms (n=791, 0.2%).

The primary schemes utilized with the corresponding boosters administered, stratified by platform, are shown in figure 2.

**Figure 2:**
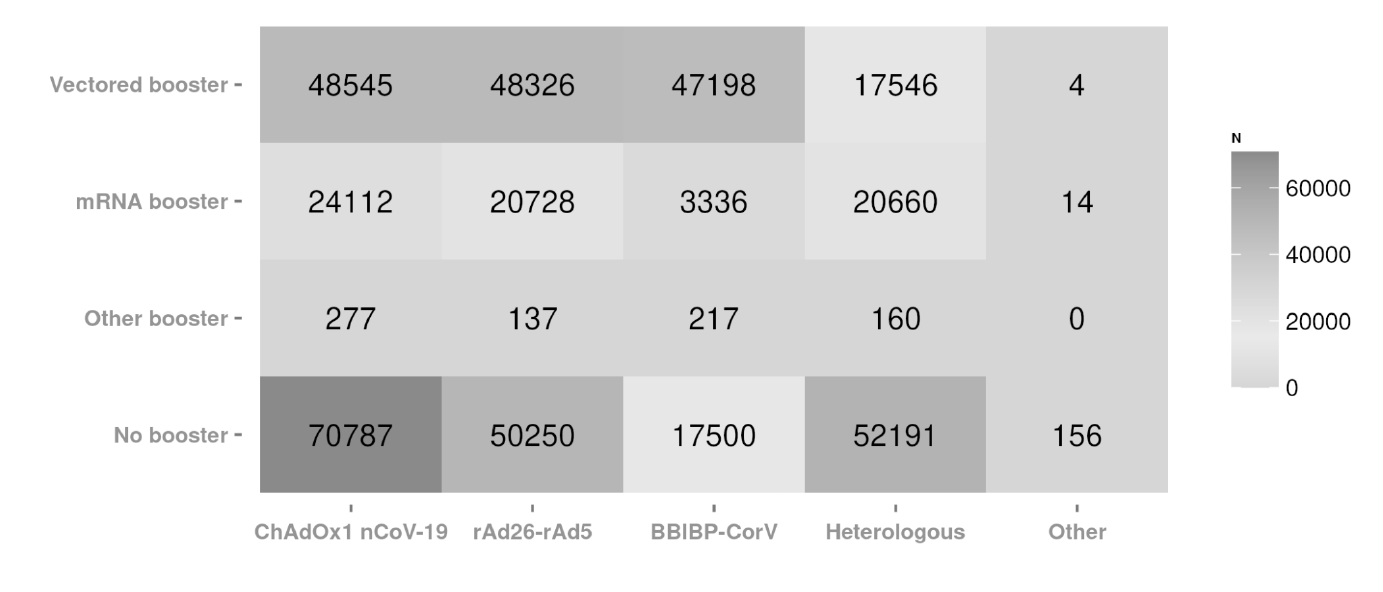
Primary schemes and boosters administered stratified by platform

Characteristics of the entire group and comparisons between two- and three-dose vaccinated subjects are shown in table 1.Briefly, compared to the two-dose subgroup, the three-dose subgroup was significantly older, had a lower proportion of males, higher proportion of subjects with comorbid conditions, and lower proportions of subjects with previously registered SARS-CoV-2 infections and previous tests performed.

**Table 1:** Characteristics of the entire group and comparisons between two- and three-dose vaccinated subjects.

|  |  | Overall | Two doses | Three doses | p |
| --- | --- | --- | --- | --- | --- |
| **n** |  | 422144 | 190,884 | 231,260 |  |
| **Age** |  | 62.19 ± 9.82 | 59.67 ± 8.90 | 64.26 ± 10.06 | <0.001 |
| **Gender** | **F** | 235,304 (55.7) | 103,704 (54.3) | 131,600 (56.9) | <0.001 |
|  | **M** | 186,840 (44.3) | 87,180 (45.7) | 99,660 (43.1) |  |
| **Comorbidities** | **No** | 394,125 (93.4) | 179,948 (94.3) | 214,177 (92.6) | <0.001 |
|  | **Yes** | 28,019 ( 6.6) | 10,936 ( 5.7) | 17,083 ( 7.4) |  |
| **Greater Buenos Aires** | **No** | 127,579 (30.2) | 54,059 (28.3) | 73,520 (31.8) | <0.001 |
|  | **Yes** | 294,565 (69.8) | 136,825 (71.7) | 157,740 (68.2) |  |
| **Previous positive SARS-CoV-2 tests** | **0** | 367,309 (87.0) | 164,285 (86.1) | 203,024 (87.8) | <0.001 |
|  | **1** | 53,244 (12.6) | 25,814 (13.5) | 27,430 (11.9) |  |
|  | **2** | 1,535 ( 0.4) | 759 ( 0.4) | 776 ( 0.3) |  |
|  | **3** | 52 ( 0.0) | 25 ( 0.0) | 27 ( 0.0) |  |
|  | **4** | 4 ( 0.0) | 1 ( 0.0) | 3 ( 0.0) |  |
| **Previous total SARS-CoV-2 tests** | **0** | 244,772 (58.0) | 110,179 (57.7) | 134,593 (58.2) | <0.001 |
|  | **1-2** | 158,275 (37.5) | 72,384 (37.9) | 85,891 (37.1) |  |
|  | **3+** | 19,097 ( 4.5) | 8,321 ( 4.4) | 10,776 ( 4.7) |  |
| **Interval after last dose in days** |  | 96 ± 59 | 149 ± 42 | 52 ± 25 | <0.001 |
| **Interval after last dose** | **15-59 days** | 156,438 (37.1) | 4,925 (2.6) | 151,513 (65.6) | <0.001 |
|  | **60-119 days** | 101,299 (24.0) | 24,882 (13.0) | 76,417 (33.0) |  |
|  | **≥120 days** | 164,407 (38.9) | 161,077 (84.4) | 3,330 (1.4) |  |
| **Events per outcome** | **Infections** | 221,933 (52.6) | 119,599 (62.7) | 102,334 (44.3) | <0.001 |
|  | **Hospitalisations** | 2,178 (0.5) | 1,341 (0.7) | 837 (0.4) | <0.001 |
|  | **Deaths** | 2,209 (0.5) | 1,420 (0.7) | 789 (0.3) | <0.001 |

Data are presented as mean ± SD and n (%)

The median time elapsed for the three-dose scheme since the application of the last dose for the <60 and ≥ 60 days subgroups was 36 (26 to 48) and 75 (66 to 89) days, respectively. The median time elapsed for the two-dose scheme since the application of the last dose for the <120 and ≥120 days subgroups was 48 (31 to 66) and 127 (123 to 132) days, respectively.

Regarding confirmed infections (table 1), there were 221,933 cases out of 422,144 (52.6%) eligible subjects; 119,599 were in the two-dose subgroup and 102,334 in the three-dose (62.7% vs 44.3%; p<0.001). Two-thousand one hundred and seventy-eight individuals were hospitalised (0.5%); 1,341 in the two-dose subgroup and 837 in the three-dose subgroup (0.7% vs 0.4%; p <0.001). A total of 2,209 (0.5%) deaths were registered; 1,420 in the two-dose subgroup and 789 in the three-dose subgroup (0.7% vs 0.3%, p <0.001).

### Matched analysis for the entire population

The matched analysis included 127,014 cases and 180,714 controls; the number of the infections, hospitalisations and deaths are shown in table 2. Regarding infections, the booster dose decreased the OR after 14-59 days of administration; after 60 days, protection dropped back to levels similar to the two-dose scheme. The booster dose also decreased the risk of hospitalisations and deaths after 15-59 days, but this protective effect persisted after administration for a median of 75 days (66 to 89) (table 2 and figure 3a). These trends were also evident when subgroups of individuals with or without comorbidities, either sex, or older than 65 years were analysed (table S1 and figure S3, a-f).

**Table 2:** Odds ratio against SARS-CoV-2 infections, hospitalizations and deaths stratified by vaccination status.

| Outcome | Vaccination status | SARS-CoV-2 positive cases | SARS-CoV-2 negative controls | Matched SARS-CoV-2 positive cases * | Matched SARS-CoV-2 negative controls | Odds Ratio | 95 % CI ** |
| --- | --- | --- | --- | --- | --- | --- | --- |
| Infection | **Total** | **221,933** | **200,211** | **127,014** | **180,714** |  |  |
|  | Two doses ineligible | 18,820 | 10,987 | 9,413 | 10,531 | 0.97 | 0.94-1.00 |
|  | Two doses eligible | 100,779 | 60,298 | 51,748 | 57,301 | 1 (ref) |  |
|  | Three doses, ≤ 60 days | 74,777 | 76,736 | 45,933 | 72,777 | 0.81 | 0.80 - 0.83 |
|  | Three doses, ≥60 days | 27,557 | 52,190 | 19,920 | 40,105 | 1.04 | 1.01-1.06 |
| Hospitalisation | **Total** | **2,178** | **200,211** | **2,149** | **9,254** |  |  |
|  | Two doses ineligible | 136 | 10,987 | 136 | 356 | 1.08 | 0.87-1.35 |
|  | Two doses eligible | 1,205 | 60,298 | 1,192 | 3,044 | 1 (ref) |  |
|  | Three doses, ≤ 60 days | 546 | 76,736 | 533 | 4,483 | 0.28 | 0.25-0.32 |
|  | Three doses, ≥60 days | 291 | 52,190 | 288 | 1,371 | 0.52 | 0.44-0.61 |
| Death | **Total** | **2,209** | **200,211** | **2,196** | **10,023** |  |  |
|  | Two doses ineligible | 136 | 10,987 | 135 | 329 | 1.09 | 0.88-1.35 |
|  | Two doses eligible | 1,284 | 60,298 | 1,275 | 3,188 | 1 (ref) |  |
|  | Three doses, ≤ 60 days | 538 | 76,736 | 537 | 5,054 | 0.25 | 0.22-0.28 |
|  | Three doses, ≥60 days | 251 | 52,190 | 249 | 1,452 | 0.38 | 0.33-0.45 |

**Figure 3:**
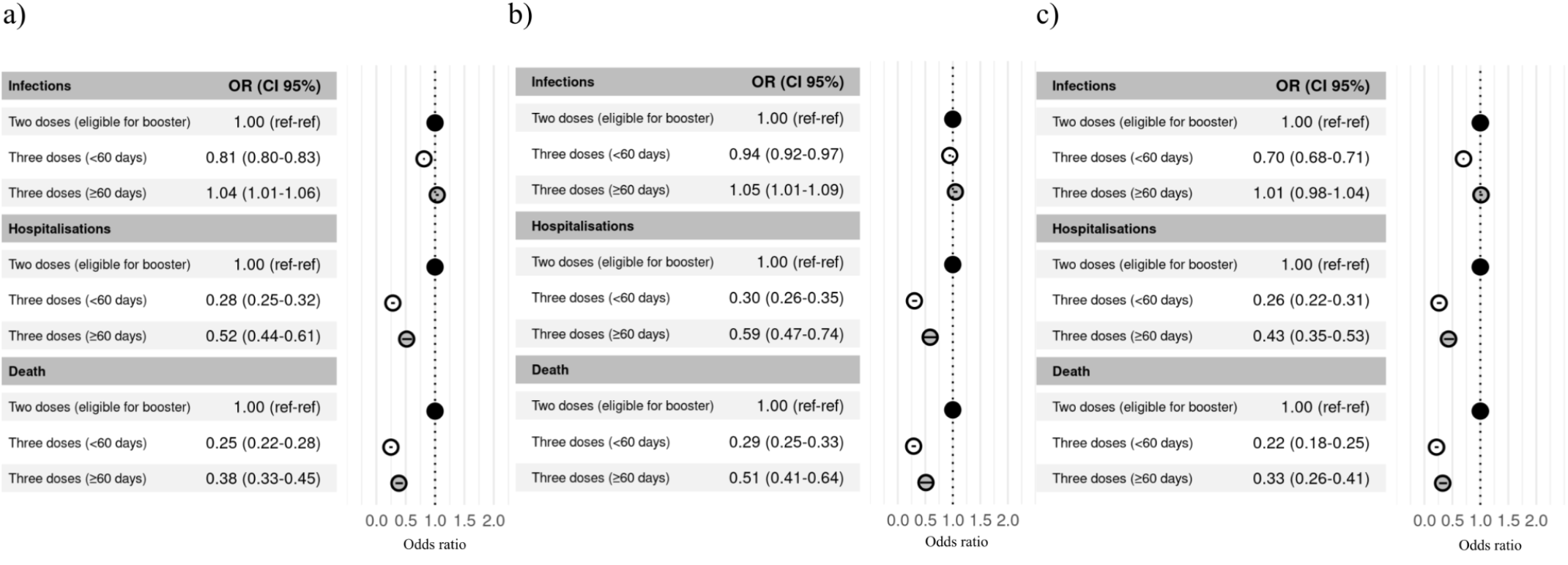
Odds ratios of boosters against confirmed SARS-CoV-2 infections, hospitalizations and death. a) All boosters b) Homologous boosters. c) Heterologous boosters.

### Protection of homologous boosters against infection, hospitalisations and death

The primary schemes with ChAdOx1 nCoV-19, rAd26-rAd5 or vectored heterologous schemes plus a vectored-vaccine booster showed similar trends regarding protection of infections, consisting in a small effect (OR 0.94; 95% CI 0.92 to 0.97), which rapidly waned (OR 1.05; 1.01 to 1.09). These schemes provided a large protection against hospitalisations and death (OR 0.30; 0.26 to 0.35 and OR 0.29; 0.25 to 0.33, respectively) but the effect of all homologous primary courses receiving a booster of a similar platform against hospitalisations and death (OR 0.59; 0.47 to 0.74 and OR 0.51; 0.41 to 0.64, respectively) waned after 60 days (figure 3 a-c and table S2).

Odds ratio for all subgroups by primary scheme and platform of booster are shown in figure 4 a-c and table S3.

**Figure 4:**
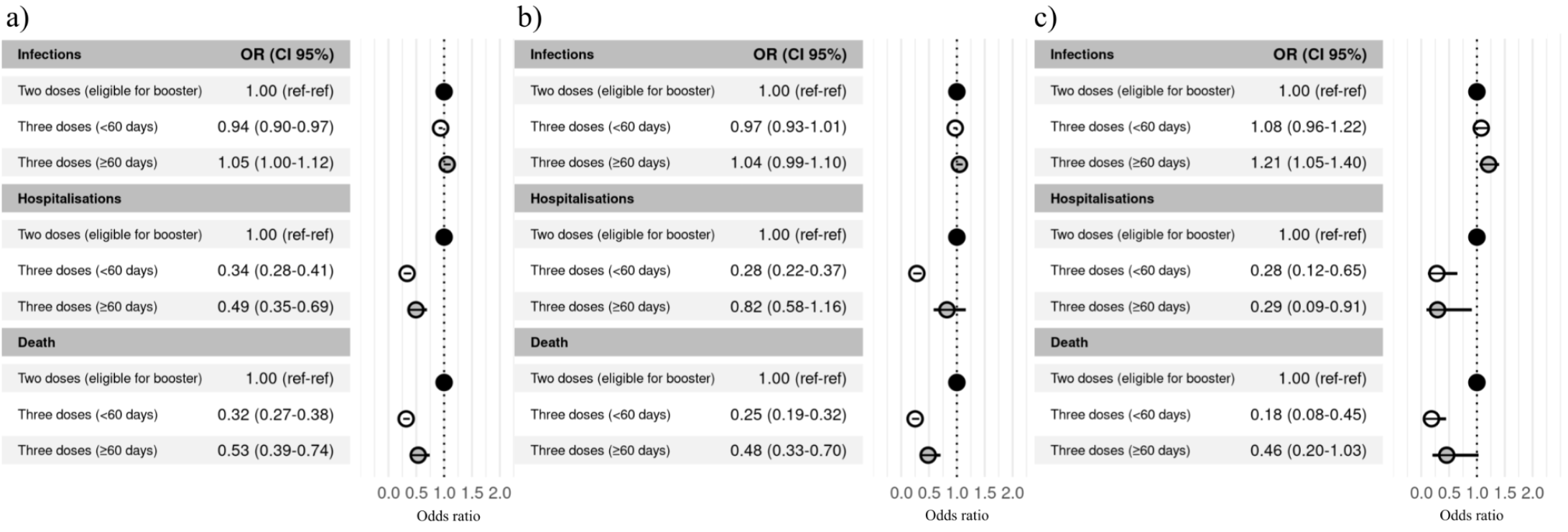
Odds ratio of homologous vectored booster against confirmed SARS-CoV-2 infections, hospitalizations and death stratified by primary scheme. a) ChAdOx1 nCoV-19 b) rAd26-rAd5 c) Vectored heterologous primary schemes.

### Protection of heterologous boosters against infection, hospitalisations and death

The primary courses with ChAdOx1 nCoV-19, rAd26-rAd5 plus a mRNA booster or with BBIBP-CorV plus mRNA or vectored booster afforded additional protection against infections (OR 0.70; 0.68 to 0.71), but an effect of waning after 60 days was evident (OR 1.01; 0.98 to 1.04) (figure 3, table S2). Notwithstanding this, there is clear protective effect against hospitalisations (OR 0.26; 0.22 to 0.31) and deaths (OR 0.22; 0.18 to 0.25) in all cases, which persists after 60 days (OR 0.43; 0.35 to 0.53 and 0.33; 0.26 to 0.41, respectively). Odds ratios for all subgroups by primary scheme and platform of booster are shown in figure 5 a-d and table S3.

**Figure 5:**
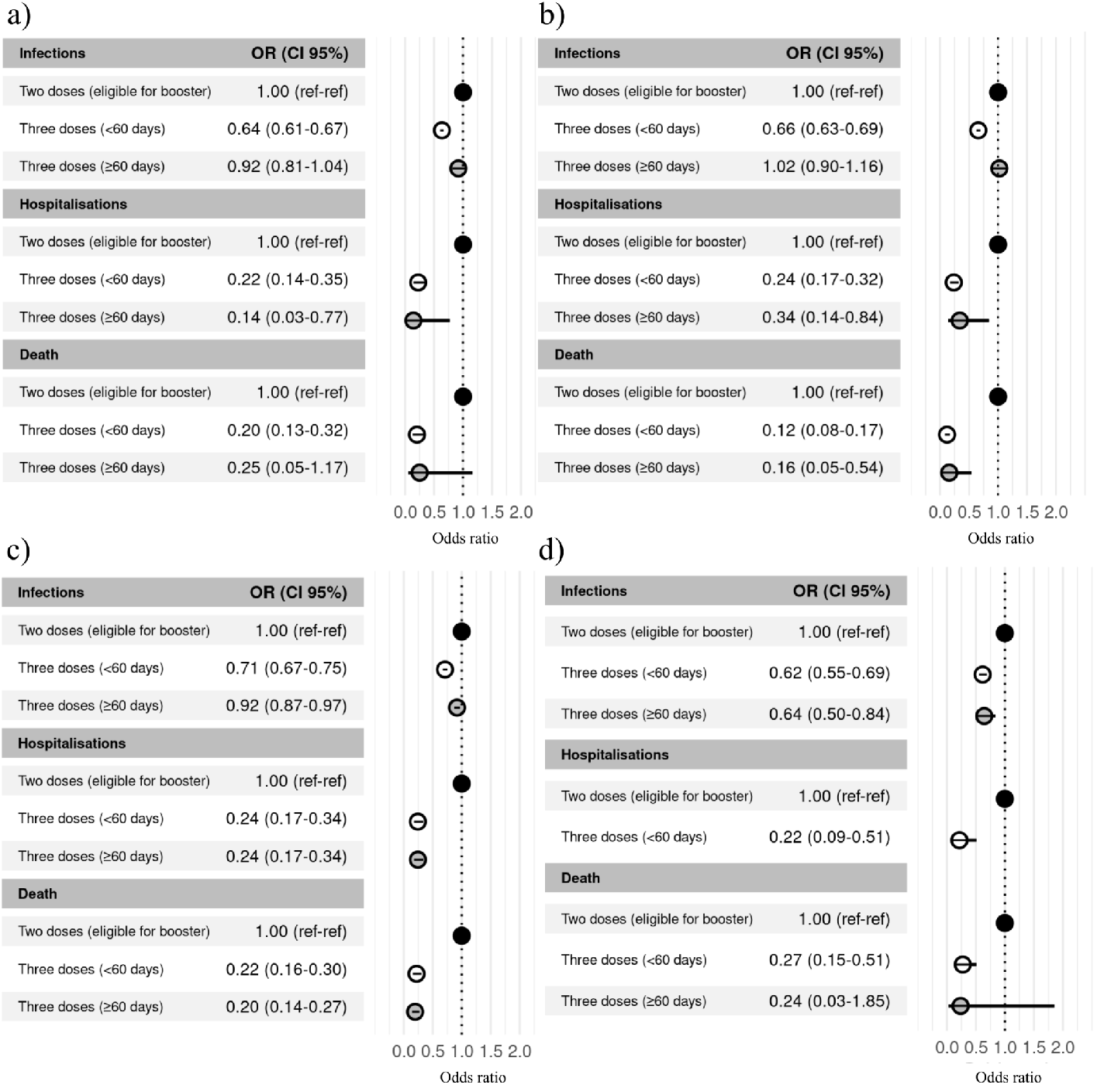
Odds ratio of heterologous boosters against confirmed SARS-CoV-2 infections, hospitalizations and death stratified by primary scheme and type of booster. a) ChAdOx1 nCoV-19 plus mRNA b) rAd26-rAd5 plus mRNA c) BBIBP-CorV plus mRNA d) BBIBP-CorV plus vectored vaccine.

After heterologous primary schemes, mRNA boosters conferred greater protection against hospitalisations and death when compared with viral vectored boosters. However, for these viral vector types of boosters the confidence intervals were wide, probably due to the small number of individuals in this category (figure 6 a-c and table S3).

**Figure 6:**
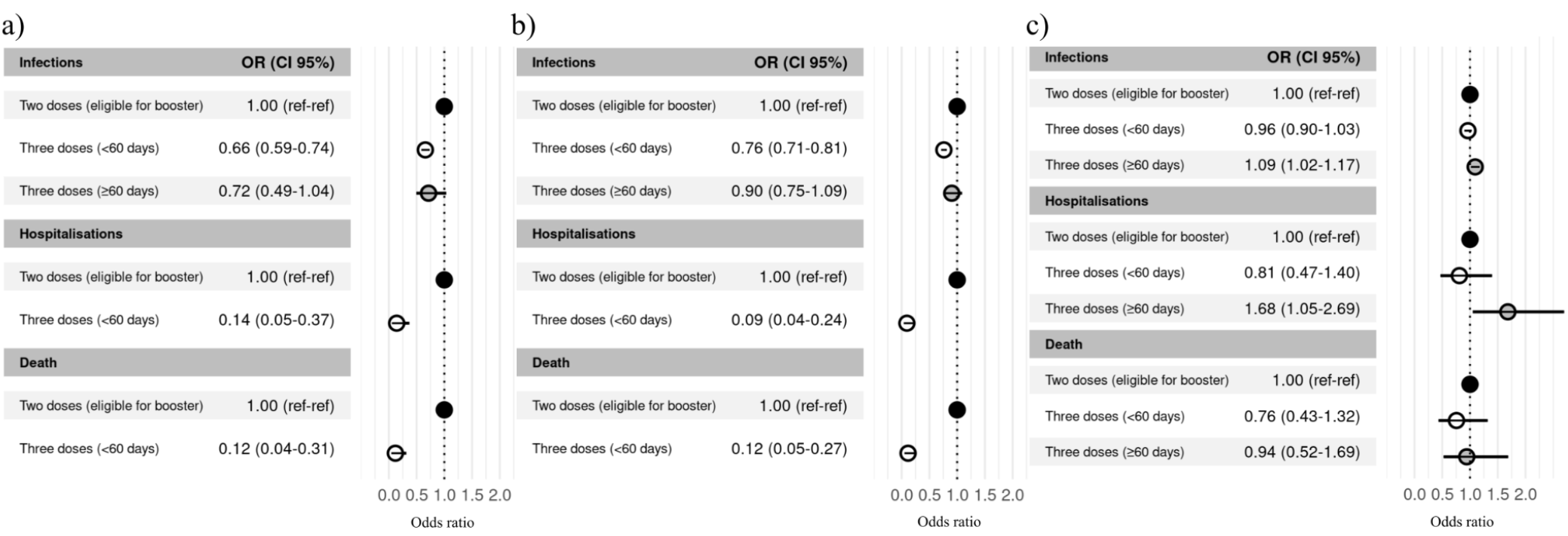
Odds ratio of heterologous boosters against confirmed SARS-CoV-2 infections, hospitalizations and death stratified by primary scheme and type of booster. a) Vectored heterologous primary schemes plus mRNA booster. b) Vectored-mRNA heterologous primary schemes plus vectored booster. c) Vectored-mRNA heterologous primary schemes plus mRNA booster

### Sensitivity analysis

We repeated this analysis stratifying the data according to two-age levels (under or over 65 years old), gender, and presence or absence of comorbidities, to assess the marginal risk reduction for each of these subgroups. We observed similar trends to those in the main analysis. In order to evaluate the effect of the time cut-off-based subgrouping in the waning analysis, models for infection, hospitalisations and death were run considering different time cut-offs. Differences observed in the ORs obtained were not statistically significant (p> 0.05, 95%CI) considering a tolerance of ± 10 days from the cut-off selected for the main analysis (60 days). To assess the uncertainty associated with the sampling, the matching process was repeated one hundred times for hospitalisations and death analysis. Thus, one hundred ORs were computed for each analysis. In both cases, as obtained ORs were within the limits of the confidence intervals reported in the main analysis for the abovementioned outcomes, it is possible to conclude that our main result is not significantly different to the result of any of these repetitions. These analyses are shown in the Supplemental Material (figures S4 and S5).

## Discussion

We examined the effect of homologous and heterologous boosters applied after different primary vaccination schemes against infection, hospitalisations and death in individuals older than 50 during omicron BA.1 predominance. Our main finding was that the administration of a heterologous booster after a primary homologous scheme produced greater beneficial effects on hospitalisations and death, which did not wane at different time points from inoculation, in comparison to homologous boosters. Similar results have been reported in previous studies.^7–10,12,13,15–18^ In addition, this study provides new information on the utilisation of the inactivated vaccine BBIBP-CorV, the viral vector vaccine rAd26-rAd5, and heterologous primary schemes, which was previously quite scarce.^19^

After realizing that ChAdOx1 nCoV-19 administration was associated with increased risk of thrombosis with thrombocytopenia syndrome, some European Union Member States embarked on a strategy of heterologous primary vaccination during the spring of 2021.^33^ Similar strategies involving heterologous vaccination in other diseases have been applied in the past.^34^ In this context, new data has emerged, acknowledging that the administration of heterologous boosters is as good as, or even better, than homologous boosters in terms of immunological response.^3,5,6–8,35^ *In vitro* studies also back this strategy, so the CDC and ECDC recommended the “mix and match”.^36,37^ In a worldwide context of primary schemes that do not include mRNA vaccines, noting that not all countries have access to them, our findings expand on additional COVID-19 vaccine combinations.

The first studies reporting heterologous boosters in real life originated in the United Kingdom during the pre-omicron period, where BNT162b2 administered after a primary scheme of ChAdOx1 nCoV-19 had 93% VE against symptomatic disease, compared to the unvaccinated.^9^ These figures are similar to the 94% VE achieved with a homologous booster of BNT162b2 after a primary scheme of BNT162b2.^10^ In Chile, after a Sinovac primary scheme, heterologous boosting with ChAdOx1 nCoV-19 or BNT162b2 was associated with higher VE than homologous boosting against symptomatic infection (90% for ChAdOx1 nCoV-19; 93% for BNT162b2; and 68% for Sinovac), hospitalisations (96%, 89%, and 75%, respectively), and intensive care unit admission (98%, 90%, and 79%), during the period of delta VOC predominance.^10^ Both studies show high VE against symptomatic infection, differently to our study in which booster administration produced a low and brief protection against this outcome. These divergent results might be ascribed to the omicron’s great capacity for immune evasion.

In our study, the effect of homologous boosting afforded small or no additional protection against confirmed infections, as was documented in Brazil and Singapore after a triple scheme with Sinovac, and in the US and Malaysia where viral vectored boosters Janssen and ChAdOx1 nCoV-19 were administered.^11–14,21,38^ Conversely, two studies from the UK, Qatar, and a systematic review using triple-mRNA reported acceptable protection against infections, but it waned after 2 or 3 months.^4,9,13,16,39^ We found that the effect of homologous boosting against hospitalisations and death was slightly lower than after the utilisation of heterologous boosting with a trend to waning after 60 days—similar to other reports.^11–13,40,41^ An exception occurred with the administration of a primary heterologous vectored scheme followed by a vectored booster, which provided a significant protection against mortality.

Concerning the use of heterologous boosters after homologous or heterologous primary schemes against infections, we observed a modest increase in protection that waned after 60 days, similar to the results described by researchers from Brazil, Scotland and the United Kingdom after the application of ChAdOx1 nCoV-19 primary schemes plus mRNA booster.^15,17,18^ In contrast, a US study with Ad26.COV2-S plus mRNA booster reported protection against infections up to 160 days.^13^ We observed, however, high additional protection against hospitalisations and death after the administration of a heterologous booster used with any primary regimen, which was maintained for a median of 75 days (IQR 66-89). Similar results were reported with heterologous boosting after the administration of primary schemes with the inactivated vaccine Sinovac and the vectored vaccines Ad26.COV2-S and ChAdOx1 nCoV-19.^13,15,17–20^ Although our results are the first to report protection in a real-life setting, they are in line with *in vitro* studies: Argentinian researchers found that a heterologous booster with ChAdOx1 nCoV-19, rAd26-rAd5, or BNT162b2 vaccines markedly increased the neutralising activity against the Omicron variant and was maintained up to 90 days in older people with primary schemes of BBIBP-CorV.^8^ Furthermore, researchers from Bahrein and Serbia reported that heterologous boosting with Pfizer-BioNTech after administering a primary scheme with rAd26-rAd5 yielded higher levels of antibodies than homologous boosting.^40,41^

To our knowledge, ours is the first study carried out in a real world setting in which primary schemes with BBIBP-CorV, rAd26-rAd5 or multiple heterologous courses were applied and then boosted with vaccines from different platforms. Additionally, we not only recorded protection against infections but also against hospitalisations and death.

### Limitations

First, we were only able to assess the effect of the booster up to a median time of 75 days due to the rapid surge and decrease of the omicron BA.1 wave; it is possible that the VE and waning effect might develop further changes over time. Nevertheless, that amount of time might be sufficient to detect patterns of change.^38–39^ Second, we could only estimate the odds ratio of a third dose relative to a second dose. The calculation of the absolute odds ratio (comparison with an unvaccinated population) was not possible given that 95% of the population older than 50 years had received at least two vaccine doses by the study period.^42–43^ Third, modification of the testing protocols during the study period might have influenced healthcare seeking behavior. The choice of a test-negative design aims to attenuate this possible bias. Fourth, misclassification cannot be completely discarded since contamination by incidental COVID-19 cases remains possible. For this reason, we only contemplated hospitalisations and death occurring within 14 days and 28 days from COVID-19 diagnosis, respectively. Fifth, viral genome sequencing was not available for most individuals; therefore, omicron BA.1 predominance periods were based on genomic surveillance data. Sixth, we only included individuals over 50; thus, our findings about VE and waning cannot be generalised to younger people. Seventh, OR estimates may be biased due to residual confounders, as in any observational study. Eighth, due to the increased burden of data entry to document infections during the short - but intense-omicron wave, it is possible that underreporting of hospitalisations occurred. Finally, given the unavailability of mRNA vaccines at the beginning of the vaccination roll-out, we could not evaluate the performance of homologous 3-dose schemes involving mRNA vaccines.

## CONCLUSIONS AND AREAS FOR FURTHER RESEARCH

This study shows that heterologous boosters provide an enhanced protection and longer effect duration against COVID-19-related hospitalisations and death in individuals older than 50 compared to homologous boosters, during Omicron predominance. The implications of our findings thus support the utilisation of different booster strategies to reach durable vaccine protection, especially in populations with primary schemes involving viral vectored or inactivated vaccines. Continuous monitoring of booster effectiveness over longer periods of time, with consideration of possible new SARS-CoV-2 variants, is key to developing the most appropriate COVID-19 vaccination strategies.

