## Supplemental material for "Protection of homologous and heterologous boosters after primary schemes of rAd26-rAd5, ChAdOx1 nCoV-19 and BBIBP-CorV during the Omicron outbreak in adults of 50 years and older in Argentina: a test-negative case-control study"

**Figure S1**: Characteristics of the COVID-19 pandemic in Argentina and the Province of Buenos Aires. A) Confirmed COVID-19 cases by epidemiological week. B) COVID-19 associated hospitalisations by epidemiological week. C) COVID-19 associated deaths by epidemiological week

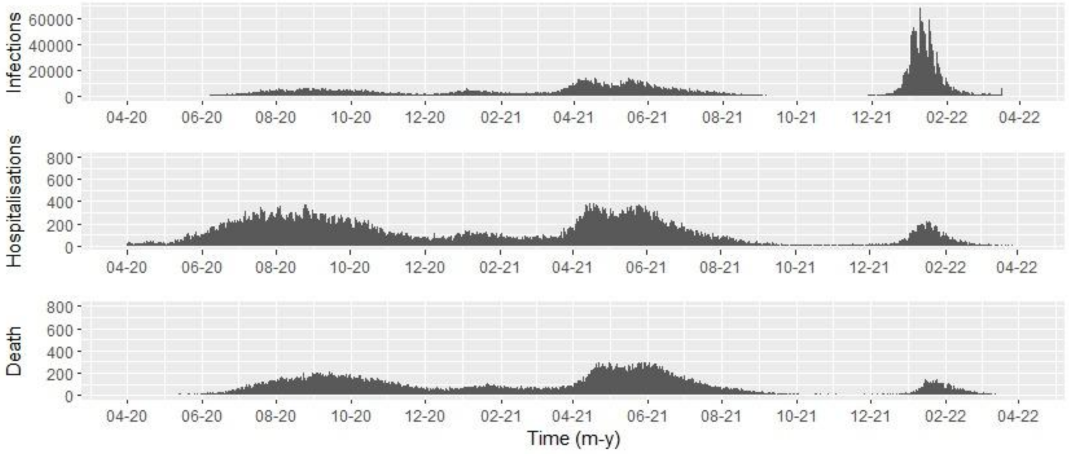


**Figure S2**: COVID-19 vaccination uptake over time in adults older than 50. From 12/27/2020 to 04/01/2022

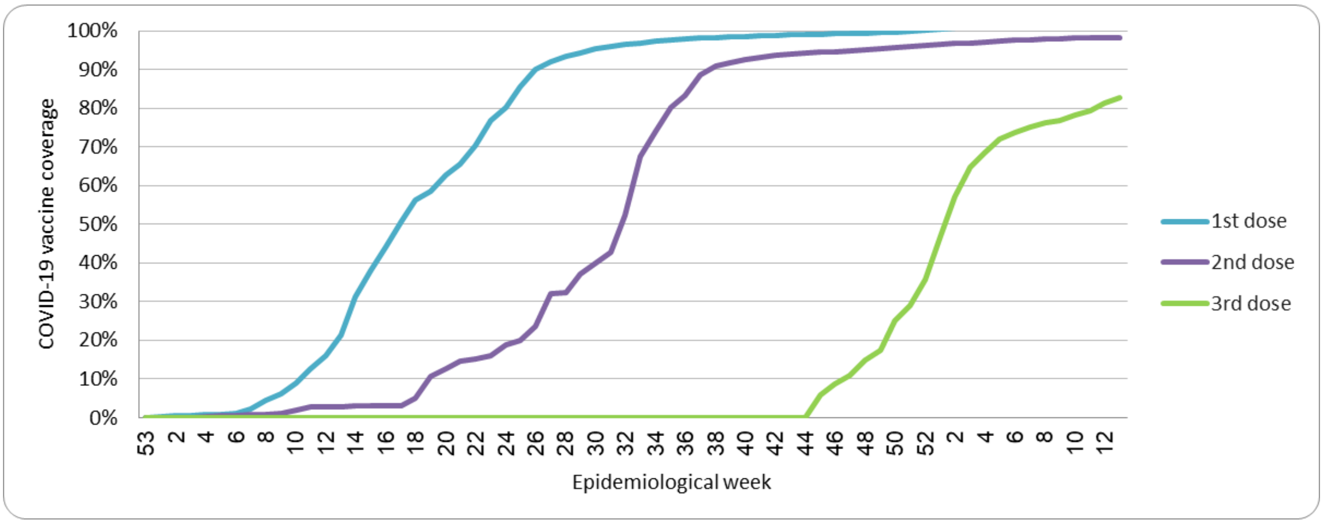

### Subgroups analysis

Binary stratified analyses according to two-age levels (under or over 65 years old), gender, and presence or absence of comorbidities to assess the marginal risk reduction for each of these subgroups. We observed similar trends to those in the main analysis.

**Figure S3**: Odds ratio of boosters against confirmed SARS-CoV-2 infections, hospitalizations and death by subgroup. a) Under 65 years old, b) Older than 65 years, c) Without comorbidities, d) With comorbidities, e) Male, f) Female.

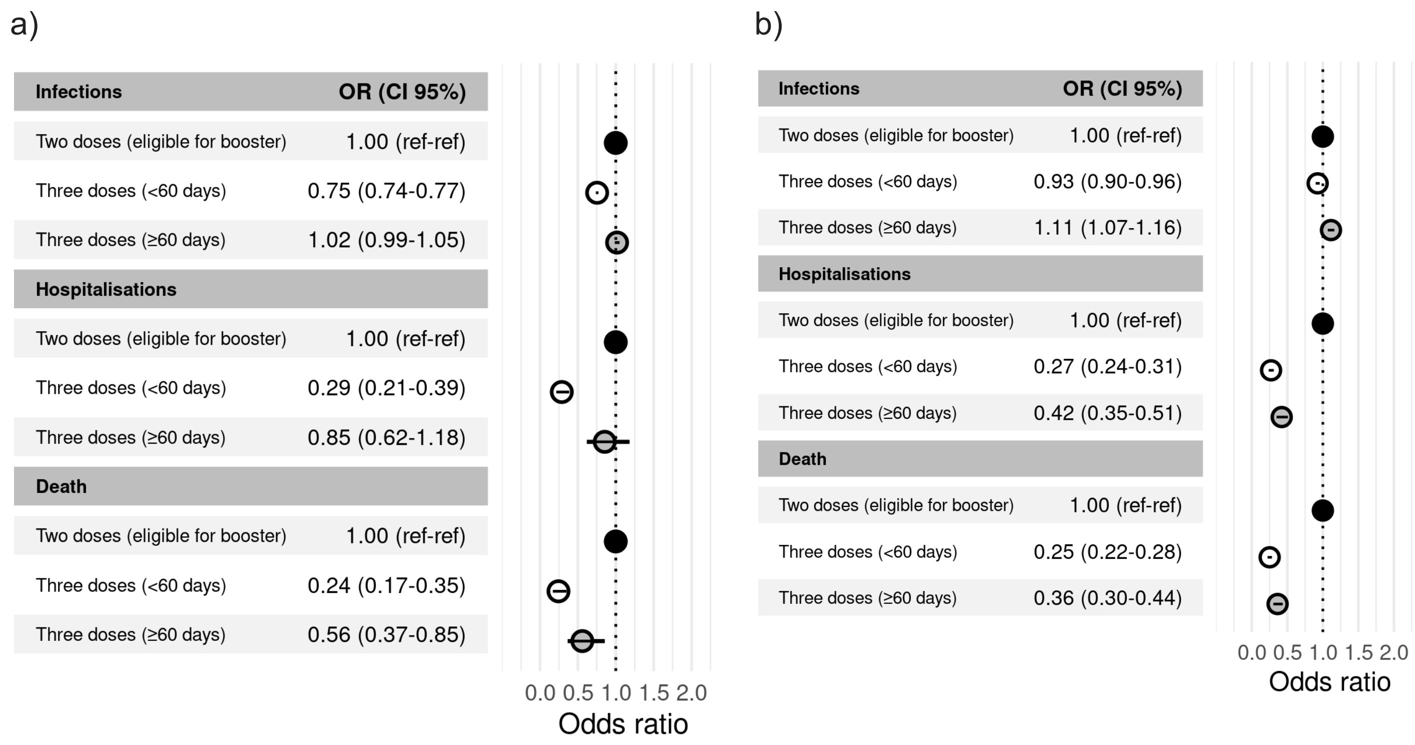

c)

| Infections | OR (CI 95%) |
|---|---|
| Two doses (eligible for booster) | 1.00 (ref-ref) |
| Three doses (<60 days) | 0.81 (0.80-0.83) |
| Three doses (≥60 days) | 1.04 (1.01-1.07) |
| **Hospitalisations** | |
| Two doses (eligible for booster) | 1.00 (ref-ref) |
| Three doses (<60 days) | 0.27 (0.23-0.32) |
| Three doses (≥60 days) | 0.39 (0.31-0.50) |
| **Death** | |
| Two doses (eligible for booster) | 1.00 (ref-ref) |
| Three doses (<60 days) | 0.26 (0.23-0.30) |
| Three doses (≥60 days) | 0.42 (0.34-0.51) |

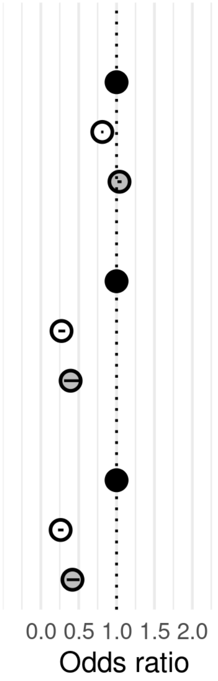


d)

| Infections | OR (CI 95%) |
|---|---|
| Two doses (eligible for booster) | 1.00 (ref-ref) |
| Three doses (<60 days) | 0.86 (0.80-0.93) |
| Three doses (≥60 days) | 1.15 (1.05-1.26) |
| **Hospitalisations** | |
| Two doses (eligible for booster) | 1.00 (ref-ref) |
| Three doses (<60 days) | 0.32 (0.27-0.38) |
| Three doses (≥60 days) | 0.65 (0.52-0.81) |
| **Death** | |
| Two doses (eligible for booster) | 1.00 (ref-ref) |
| Three doses (<60 days) | 0.24 (0.20-0.30) |
| Three doses (≥60 days) | 0.39 (0.29-0.53) |

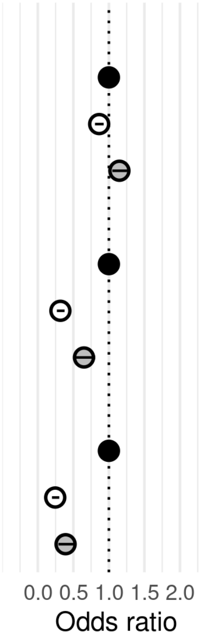


e)

| Infections | OR (CI 95%) |
|---|---|
| Two doses (eligible for booster) | 1.00 (ref-ref) |
| Three doses (<60 days) | 0.80 (0.77-0.82) |
| Three doses (≥60 days) | 1.01 (0.98-1.05) |
| **Hospitalisations** | |
| Two doses (eligible for booster) | 1.00 (ref-ref) |
| Three doses (<60 days) | 0.26 (0.22-0.31) |
| Three doses (≥60 days) | 0.51 (0.41-0.64) |
| **Death** | |
| Two doses (eligible for booster) | 1.00 (ref-ref) |
| Three doses (<60 days) | 0.24 (0.20-0.28) |
| Three doses (≥60 days) | 0.43 (0.35-0.54) |

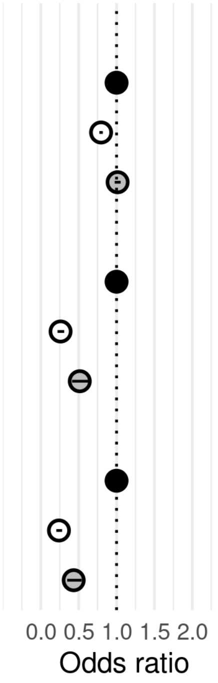


f)

| Infections | OR (CI 95%) |
|---|---|
| Two doses (eligible for booster) | 1.00 (ref-ref) |
| Three doses (<60 days) | 0.81 (0.79-0.83) |
| Three doses (≥60 days) | 1.06 (1.02-1.09) |
| **Hospitalisations** | |
| Two doses (eligible for booster) | 1.00 (ref-ref) |
| Three doses (<60 days) | 0.31 (0.27-0.37) |
| Three doses (≥60 days) | 0.50 (0.39-0.64) |
| **Death** | |
| Two doses (eligible for booster) | 1.00 (ref-ref) |
| Three doses (<60 days) | 0.26 (0.22-0.31) |
| Three doses (≥60 days) | 0.32 (0.25-0.42) |

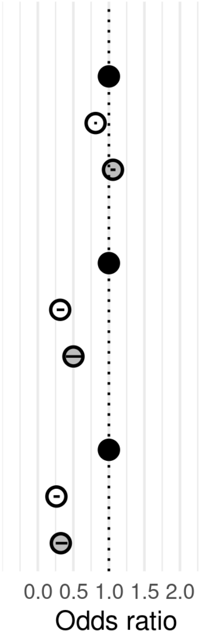

**Figure S4: Uncertainty associated with the process of matching cases and controls.** In order to assess the uncertainty associated with the process of matching cases and controls, the matching and modeling process was repeated for a total of 100 times per outcome. Due to computational intensiveness, this sensitivity analysis was only performed for outcomes associated with severe disease. For each of these matched sets, a conditional logistic regression model was fitted with each vaccination status odds ratio analyzed. Every OR estimated is shown in the figure above. Horizontal lines show the results shown in the main analysis, with its IC95 confidence interval represented with dashed lines.

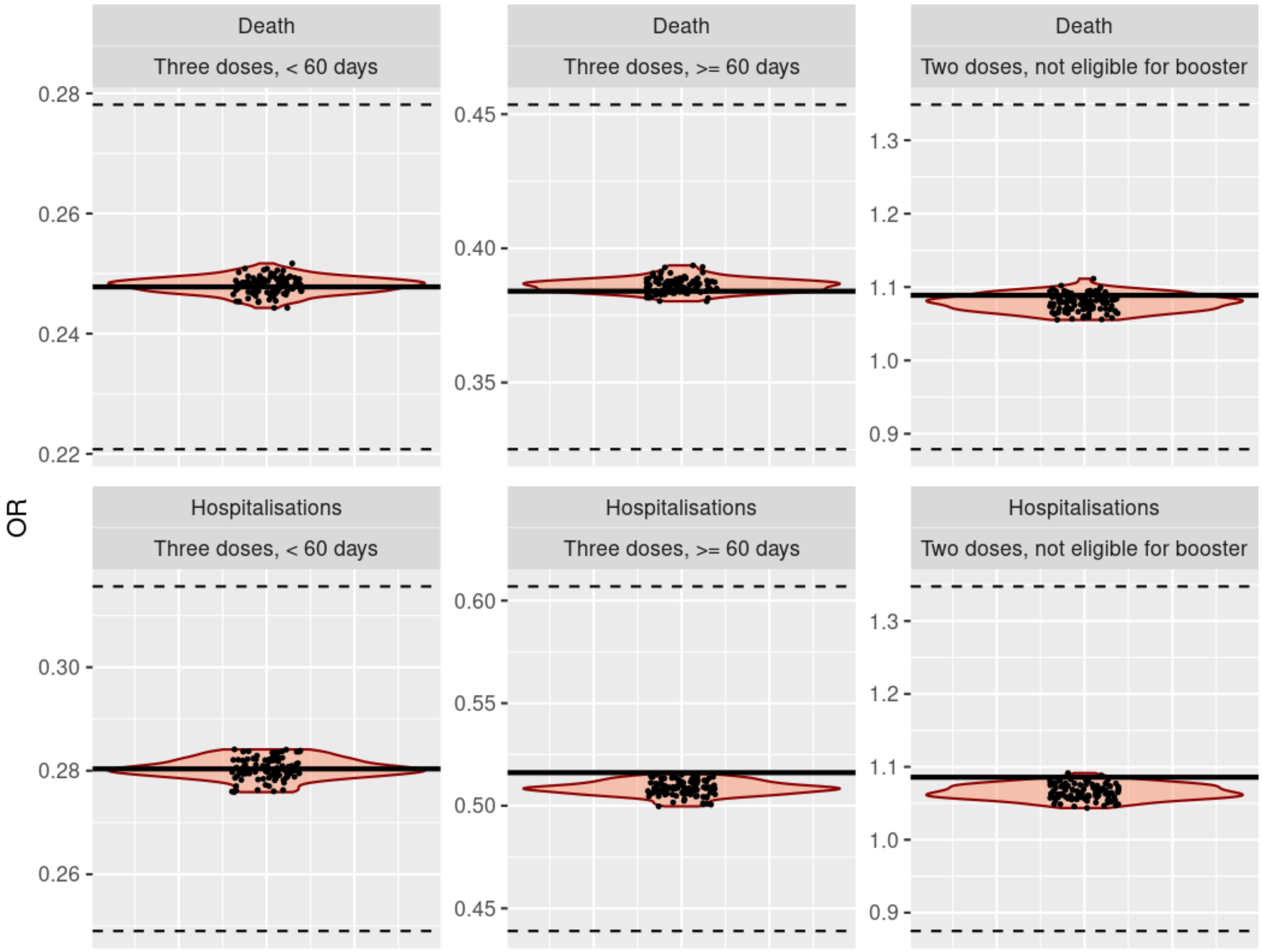

**Figure S5: Interval considered for dividing three dose vaccinated individuals**

In order to assess the change in the effect of the three doses vaccine by the temporal interval (time between vaccination and onset of symptoms) we estimated this effect in each outcome taking different intervals. For each outcome a conditional logistic regression model was fitted for each interval and the vaccination status odds ratio was analyzed. We summarize this information in the graphics shown below. For each outcome we plot the change in the OR according to the interval taken. The vertical bars at each point represent the IC95 confidence interval.

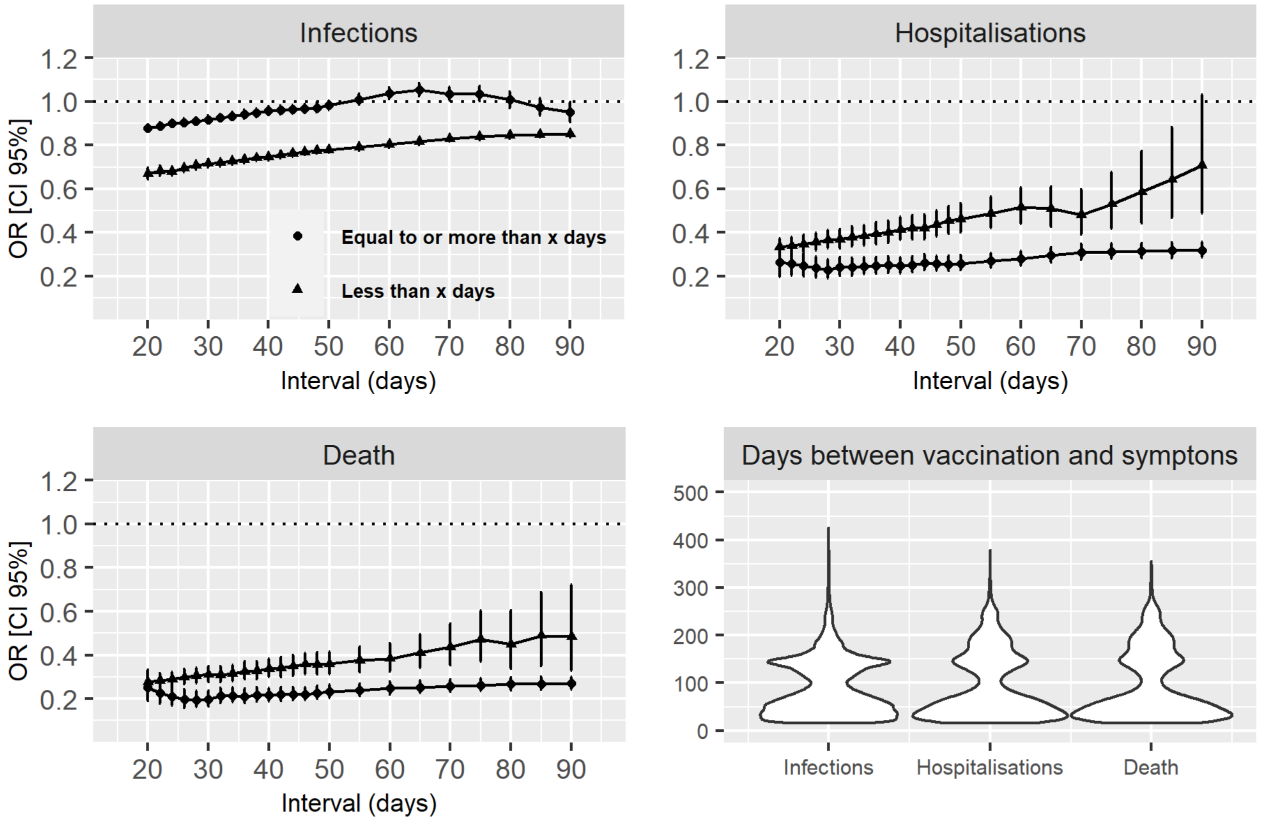

**Table S1**: Odds ratio of booster against confirmed SARS-CoV-2 infections, hospitalizations and death by subgroup. a) Under 65 years old, b) Over 65 years old, c) Without comorbidities, d) With comorbidities, e) Male, f) Female

| Subgroup | Vaccination status | Outcome | Matched SARS-CoV-2 positive cases | Matched SARS-CoV-2 negative controls | Odds ratio | CILow | CIHigh |
|---|---|---|---|---|---|---|---|
| 50-65 years old | Two doses elegible | Infections | 38382 | 41196 | 1,00 | Ref | Ref |
| | Two doses ineligible | | 7891 | 8673 | 0,96 | 0,93 | 1,00 |
| | Three doses ≥60 days | | 11433 | 21943 | 1,02 | 0,99 | 1,05 |
| | Three doses, ≤ 60 days | | 24276 | 42408 | 0,75 | 0,74 | 0,77 |
| | Two doses elegible | Hospitalisations | 240 | 865 | 1,00 | Ref | Ref |
| | Two doses ineligible | | 48 | 159 | 1,10 | 0,77 | 1,58 |
| | Three doses ≥60 days | | 73 | 286 | 0,85 | 0,62 | 1,18 |
| | Three doses, ≤ 60 days | | 69 | 798 | 0,29 | 0,21 | 0,39 |
| | Two doses elegible | Death | 169 | 577 | 1,00 | Ref | Ref |
| | Two doses ineligible | | 38 | 95 | 1,37 | 0,90 | 2,07 |
| | Three doses ≥60 days | | 35 | 182 | 0,56 | 0,37 | 0,85 |
| | Three doses, ≤ 60 days | | 43 | 553 | 0,24 | 0,17 | 0,35 |
| 65+ years old | Two doses elegible | Infections | 13164 | 15931 | 1,00 | Ref | Ref |
| | Two doses ineligible | | 1503 | 1852 | 0,99 | 0,92 | 1,07 |
| | Three doses ≥60 days | | 8476 | 17808 | 1,11 | 1,07 | 1,16 |
| | Three doses, ≤ 60 days | | 21556 | 30120 | 0,93 | 0,90 | 0,96 |
| | Two doses elegible | Hospitalisations | 942 | 2117 | 1,00 | Ref | Ref |
| | Two doses ineligible | | 86 | 198 | 1,03 | 0,78 | 1,36 |
| | Three doses ≥60 days | | 212 | 1075 | 0,42 | 0,35 | 0,51 |
| | Three doses, ≤ 60 days | | 464 | 3594 | 0,27 | 0,24 | 0,31 |
| | Two doses elegible | Death | 1097 | 2582 | 1,00 | Ref | Ref |
| | Two doses ineligible | | 97 | 241 | 0,93 | 0,72 | 1,20 |
| | Three doses ≥60 days | | 213 | 1249 | 0,36 | 0,30 | 0,44 |
| | Three doses, ≤ 60 days | | 490 | 4381 | 0,25 | 0,22 | 0,28 |
| Female | Two doses elegible | Infections | 29469 | 32322 | 1,00 | Ref | Ref |
| | Two doses ineligible | | 5466 | 5887 | 1,00 | 0,96 | 1,04 |
| | Three doses ≥60 days | | 11776 | 22818 | 1,06 | 1,02 | 1,09 |
| | Three doses, ≤ 60 days | | 27089 | 41803 | 0,81 | 0,79 | 0,83 |
| | Two doses elegible | Hospitalisations | 555 | 1525 | 1,00 | Ref | Ref |
| | Two doses ineligible | | 54 | 164 | 1,01 | 0,73 | 1,41 |
| | Three doses ≥60 days | | 128 | 642 | 0,50 | 0,39 | 0,64 |
| | Three doses, ≤ 60 days | | 263 | 2139 | 0,31 | 0,27 | 0,37 |
| | Two doses elegible | Death | 615 | 1580 | 1,00 | Ref | Ref |
| | Two doses ineligible | | 50 | 158 | 0,86 | 0,62 | 1,21 |
| | Three doses ≥60 days | | 101 | 680 | 0,32 | 0,25 | 0,42 |
| | Three doses, ≤ 60 days | | 261 | 2378 | 0,26 | 0,22 | 0,31 |
| Male | Two doses elegible | Infections | 22299 | 24991 | 1,00 | Ref | Ref |

| | | | | | | | |
|---|---|---|---|---|---|---|---|
| | Two doses ineligible | | 4054 | 4654 | 0,96 | 0,92 | 1,01 |
| | Three doses ≥60 days | | 8237 | 17261 | 1,01 | 0,98 | 1,05 |
| | Three doses, ≤ 60 days | | 18623 | 30952 | 0,80 | 0,77 | 0,82 |
| | Two doses elegible | Hospitalisations | 636 | 1563 | 1,00 | Ref | Ref |
| | Two doses ineligible | | 82 | 183 | 1,17 | 0,87 | 1,56 |
| | Three doses ≥60 days | | 160 | 747 | 0,51 | 0,41 | 0,64 |
| | Three doses, ≤ 60 days | | 274 | 2294 | 0,26 | 0,22 | 0,31 |
| | Two doses elegible | Death | 661 | 1604 | 1,00 | Ref | Ref |
| | Two doses ineligible | | 85 | 165 | 1,33 | 1,00 | 1,76 |
| | Three doses ≥60 days | | 148 | 779 | 0,43 | 0,35 | 0,54 |
| | Three doses, ≤ 60 days | | 275 | 2675 | 0,24 | 0,20 | 0,28 |
| With comorbidities | Two doses elegible | Infections | 2961 | 3336 | 1,00 | Ref | Ref |
| | Two doses ineligible | | 420 | 463 | 1,00 | 0,87 | 1,16 |
| | Three doses ≥60 days | | 1806 | 3077 | 1,15 | 1,05 | 1,26 |
| | Three doses, ≤ 60 days | | 3343 | 4743 | 0,86 | 0,80 | 0,93 |
| | Two doses elegible | Hospitalisations | 588 | 1341 | 1,00 | Ref | Ref |
| | Two doses ineligible | | 58 | 150 | 0,96 | 0,69 | 1,33 |
| | Three doses ≥60 days | | 179 | 693 | 0,65 | 0,52 | 0,81 |
| | Three doses, ≤ 60 days | | 285 | 1872 | 0,32 | 0,27 | 0,38 |
| | Two doses elegible | Death | 449 | 963 | 1,00 | Ref | Ref |
| | Two doses ineligible | | 48 | 102 | 1,13 | 0,78 | 1,66 |
| | Three doses ≥60 days | | 77 | 408 | 0,39 | 0,29 | 0,53 |
| | Three doses, ≤ 60 days | | 165 | 1350 | 0,24 | 0,20 | 0,30 |
| Without comorbidities | Two doses elegible | Infections | 48629 | 53969 | 1,00 | Ref | Ref |
| | Two doses ineligible | | 9013 | 10074 | 0,98 | 0,95 | 1,01 |
| | Three doses ≥60 days | | 18244 | 36992 | 1,04 | 1,01 | 1,07 |
| | Three doses, ≤ 60 days | | 42692 | 68093 | 0,81 | 0,80 | 0,83 |
| | Two doses elegible | Hospitalisations | 597 | 1760 | 1,00 | Ref | Ref |
| | Two doses ineligible | | 78 | 198 | 1,22 | 0,91 | 1,63 |
| | Three doses ≥60 days | | 109 | 690 | 0,39 | 0,31 | 0,50 |
| | Three doses, ≤ 60 days | | 255 | 2540 | 0,27 | 0,23 | 0,32 |
| | Two doses elegible | Death | 827 | 2286 | 1,00 | Ref | Ref |
| | Two doses ineligible | | 87 | 230 | 1,04 | 0,80 | 1,36 |
| | Three doses ≥60 days | | 172 | 1006 | 0,42 | 0,34 | 0,51 |
| | Three doses, ≤ 60 days | | 369 | 3673 | 0,26 | 0,23 | 0,30 |

* Matching process used the nearest neighbor (1nn) matching, with up to five controls per case, based on age, gender, number of positive tests in the past, site of residence, presence or absence of comorbidities and number of previous tests.
* Results with IC 3 were not shown.

**Table S2**: Odds ratio of booster against confirmed SARS-CoV-2 infections, hospitalizations and deaths stratified by booster strategy.

| **Booster strategy** | **Vaccination status** | **Outcome** | **Matched SARS-CoV-2 positive cases** | **Matched SARS-CoV-2 negative controls** | **Odds ratio** | **CILow** | **CIHigh** |
|---|---|---|---|---|---|---|---|
| Homologous | Two doses elegible | Infections | 33787 | 39007 | 1.00 | Ref | Ref |
| | Two doses ineligible | | 7196 | 8332 | 0.93 | 0.90 | 0.97 |
| | Three doses ≥60 days | | 7819 | 17052 | 1.05 | 1.01 | 1.09 |
| | Three doses, ≤ 60 days | | 22677 | 27391 | 0.94 | 0.92 | 0.97 |
| | Two doses elegible | Hospitalisations | 855 | 2305 | 1.00 | Ref | Ref |
| | Two doses ineligible | | 95 | 299 | 0.95 | 0.73 | 1.22 |
| | Three doses ≥60 days | | 144 | 671 | 0.59 | 0.47 | 0.74 |
| | Three doses, ≤ 60 days | | 316 | 2529 | 0.30 | 0.26 | 0.35 |
| | Two doses elegible | Death | 926 | 2559 | 1.00 | Ref | Ref |
| | Two doses ineligible | | 98 | 280 | 1.04 | 0.81 | 1.34 |
| | Three doses ≥60 days | | 126 | 682 | 0.51 | 0.41 | 0.64 |
| | Three doses, ≤ 60 days | | 335 | 3054 | 0.29 | 0.25 | 0.33 |
| Heterologous | Two doses elegible | Infections | 51785 | 56930 | 1.00 | Ref | Ref |
| | Two doses ineligible | | 9361 | 10497 | 0.97 | 0.94 | 1.00 |
| | Three doses ≥60 days | | 12060 | 22699 | 1.01 | 0.98 | 1.04 |
| | Three doses, ≤ 60 days | | 21849 | 44868 | 0.70 | 0.68 | 0.71 |
| | Two doses elegible | Hospitalisations | 1177 | 3469 | 1.00 | Ref | Ref |
| | Two doses ineligible | | 136 | 397 | 1.08 | 0.87 | 1.33 |
| | Three doses ≥60 days | | 142 | 869 | 0.43 | 0.35 | 0.53 |
| | Three doses, ≤ 60 days | | 216 | 2292 | 0.26 | 0.22 | 0.31 |
| | Two doses | Death | 1270 | 3726 | 1.00 | Ref | Ref |

| | | | | | | | |
|---|---|---|---|---|---|---|---|
| | elegible | | | | | | |
| | Two doses ineligible | | 135 | 383 | 1.08 | 0.87 | 1.33 |
| | Three doses ≥60 days | | 121 | 945 | 0.33 | 0.26 | 0.41 |
| | Three doses, ≤ 60 days | | 197 | 2551 | 0.22 | 0.18 | 0.25 |

* Matching process used the nearest neighbour (1nn) matching, with up to five controls per case, based on age, gender, number of positive tests in the past, site of residence, presence or absence of comorbidities and number of previous tests.

**Table S3**: Odds ratio of booster against confirmed SARS-CoV-2 infections, hospitalizations and deaths stratified by primary scheme and type of platform.

| **Primary scheme** | **Platform of booster** | **Vaccination status** | **Outcome** | **Matched SARS-CoV-2 positive cases** | **Matched SARS-CoV-2 negative controls** | **Odds ratio †** | **CILow** | **CIHigh** |
|---|---|---|---|---|---|---|---|---|
| ChAdOx1 nCoV-19 | mRNA | Two doses elegible | Infections | 19448 | 21049 | 1.00 | Ref | Ref |
| | | Two doses ineligible | | 1903 | 2224 | 0.90 | 0.84 | 0.96 |
| | | Three doses ≥60 days | | 574 | 2292 | 0.92 | 0.81 | 1.04 |
| | | Three doses, ≤ 60 days | | 4994 | 11682 | 0.64 | 0.61 | 0.67 |
| | | Two doses elegible | Hospitalisations | 264 | 861 | 1.00 | Ref | Ref |
| | | Two doses ineligible | | 19 | 65 | 0.95 | 0.54 | 1.67 |
| | | Three doses ≥60 days | | 3 | 19 | 0.14 | 0.03 | 0.77 |
| | | Three doses, ≤ 60 days | | 27 | 329 | 0.22 | 0.14 | 0.35 |
| | | Two doses elegible | Death | 285 | 888 | 1.00 | Ref | Ref |
| | | Two doses ineligible | | 22 | 74 | 0.92 | 0.55 | 1.54 |
| | | Three doses ≥60 days | | 2 | 19 | 0.25 | 0.05 | 1.17 |
| | | Three doses, ≤ 60 days | | 26 | 356 | 0.20 | 0.13 | 0.32 |
| ChAdOx1 nCoV-19 | Vectored | Two doses elegible | Infections | 18745 | 21373 | 1.00 | Ref | Ref |
| | | Two doses ineligible | | 1818 | 2251 | 0.88 | 0.82 | 0.94 |
| | | Three doses ≥60 days | | 3666 | 8035 | 1.04 | 0.99 | 1.10 |
| | | Three doses, ≤ 60 days | | 10574 | 12613 | 0.97 | 0.93 | 1.01 |
| | | Two doses elegible | Hospitalisations | 273 | 770 | 1.00 | Ref | Ref |
| | | Two doses ineligible | | 20 | 60 | 0.91 | 0.52 | 1.57 |
| | | Three doses ≥60 days | | 68 | 243 | 0.82 | 0.58 | 1.16 |

| | | | | | | | | |
|---|---|---|---|---|---|---|---|---|
| | mRNA | Three doses, ≤ 60 days | | 106 | 925 | 0.28 | 0.22 | 0.37 |
| | | Two doses elegible | Death | 294 | 752 | 1.00 | Ref | Ref |
| | | Two doses ineligible | | 21 | 57 | 0.94 | 0.55 | 1.61 |
| | | Three doses ≥60 days | | 52 | 258 | 0.48 | 0.33 | 0.70 |
| | | Three doses, ≤ 60 days | | 115 | 1021 | 0.25 | 0.19 | 0.32 |
| BBIBP-CorV | mRNA | Two doses elegible | Infections | 4040 | 4759 | 1.00 | Ref | Ref |
| | | Two doses ineligible | | 1015 | 1264 | 0.94 | 0.86 | 1.03 |
| | | Three doses ≥60 days | | 96 | 364 | 0.64 | 0.50 | 0.84 |
| | | Three doses, ≤ 60 days | | 585 | 1386 | 0.62 | 0.55 | 0.69 |
| | | Two doses elegible | Hospitalisations | 144 | 437 | 1.00 | Ref | Ref |
| | | Two doses ineligible | | 23 | 79 | 1.04 | 0.61 | 1.76 |
| | | Three doses ≥60 days | | 0 | 9 | NA | NA | NA |
| | | Three doses, ≤ 60 days | | 9 | 89 | 0.22 | 0.09 | 0.51 |
| | | Two doses elegible | Death | 184 | 552 | 1.00 | Ref | Ref |
| | | Two doses ineligible | | 25 | 102 | 0.87 | 0.54 | 1.41 |
| | | Three doses ≥60 days | | 1 | 15 | 0.24 | 0.03 | 1.85 |
| | | Three doses, ≤ 60 days | | 13 | 148 | 0.27 | 0.15 | 0.51 |
| BBIBP-CorV | Vectored | Two doses elegible | Infections | 4587 | 4984 | 1.00 | Ref | Ref |
| | | Two doses ineligible | | 1218 | 1333 | 0.98 | 0.89 | 1.07 |
| | | Three doses ≥60 days | | 8021 | 13002 | 0.92 | 0.87 | 0.97 |
| | | Three doses, ≤ 60 days | | 6244 | 7615 | 0.71 | 0.67 | 0.75 |
| | | Two doses elegible | Hospitalisations | 168 | 285 | 1.00 | Ref | Ref |
| | | Two doses ineligible | | 27 | 41 | 1.18 | 0.66 | 2.11 |
| | | Three doses ≥60 days | | 91 | 593 | 0.24 | 0.17 | 0.34 |
| | | Three doses, ≤ 60 days | | 78 | 533 | 0.24 | 0.17 | 0.34 |
| | | Two doses elegible | Death | 201 | 343 | 1.00 | Ref | Ref |
| | | Two doses ineligible | | 29 | 64 | 0.72 | 0.44 | 1.20 |
| | | Three doses ≥60 days | | 89 | 687 | 0.20 | 0.14 | 0.27 |
| | | Three doses, ≤ 60 days | | 79 | 587 | 0.22 | 0.16 | 0.30 |
| rAd26-rAd5 | mRNA | Two doses elegible | Infections | 11529 | 13255 | 1.00 | Ref | Ref |
| | | Two doses ineligible | | 4525 | 4715 | 1.04 | 0.98 | 1.10 |
| | | Three doses ≥60 days | | 514 | 1832 | 1.02 | 0.90 | 1.16 |

| | | | | | | | | |
|---|---|---|---|---|---|---|---|---|
| | | Three doses, ≤ 60 days | | 4207 | 9414 | 0.66 | 0.63 | 0.69 |
| | | Two doses elegible | Hospitalisations | 440 | 1293 | 1.00 | Ref | Ref |
| | | Two doses ineligible | | 62 | 175 | 1.31 | 0.93 | 1.84 |
| | | Three doses ≥60 days | | 7 | 52 | 0.34 | 0.14 | 0.84 |
| | | Three doses, ≤ 60 days | | 62 | 690 | 0.24 | 0.17 | 0.32 |
| | | Two doses elegible | Death | 499 | 1477 | 1.00 | Ref | Ref |
| | | Two doses ineligible | | 53 | 157 | 1.21 | 0.85 | 1.71 |
| | | Three doses ≥60 days | | 4 | 44 | 0.16 | 0.05 | 0.54 |
| | | Three doses, ≤ 60 days | | 40 | 812 | 0.12 | 0.08 | 0.17 |
| rAd26-rAd5 | Vectored | Two doses elegible | Infections | 11278 | 13568 | 1.00 | Ref | Ref |
| | | Two doses ineligible | | 4603 | 4764 | 1.06 | 1.01 | 1.12 |
| | | Three doses ≥60 days | | 3565 | 7874 | 1.05 | 1.00 | 1.12 |
| | | Three doses, ≤ 60 days | | 11199 | 13473 | 0.94 | 0.90 | 0.97 |
| | | Two doses elegible | Hospitalisations | 451 | 1152 | 1.00 | Ref | Ref |
| | | Two doses ineligible | | 64 | 168 | 1.23 | 0.87 | 1.72 |
| | | Three doses ≥60 days | | 69 | 361 | 0.49 | 0.35 | 0.69 |
| | | Three doses, ≤ 60 days | | 193 | 1359 | 0.34 | 0.28 | 0.41 |
| | | Two doses elegible | Death | 507 | 1396 | 1.00 | Ref | Ref |
| | | Two doses ineligible | | 54 | 150 | 1.06 | 0.75 | 1.49 |
| | | Three doses ≥60 days | | 64 | 336 | 0.53 | 0.39 | 0.74 |
| | | Three doses, ≤ 60 days | | 212 | 1718 | 0.32 | 0.27 | 0.38 |
| Vectored heterologous | mRNA | Two doses elegible | Infections | 3179 | 3502 | 1.00 | Ref | Ref |
| | | Two doses ineligible | | 1058 | 1195 | 0.94 | 0.84 | 1.04 |
| | | Three doses ≥60 days | | 71 | 299 | 0.72 | 0.49 | 1.04 |
| | | Three doses, ≤ 60 days | | 945 | 2154 | 0.66 | 0.59 | 0.74 |
| | | Two doses elegible | Hospitalisations | 91 | 266 | 1.00 | Ref | Ref |
| | | Two doses ineligible | | 8 | 46 | 0.47 | 0.20 | 1.09 |
| | | Three doses ≥60 days | | 0 | 2 | NA | NA | NA |
| | | Three doses, ≤ 60 days | | 6 | 93 | 0.14 | 0.05 | 0.37 |
| | | Two doses elegible | Death | 92 | 273 | 1.00 | Ref | Ref |
| | | Two doses ineligible | | 14 | 54 | 0.60 | 0.29 | 1.25 |
| | | Three doses ≥60 days | | 0 | 2 | NA | NA | NA |

| | | | | | | | | |
|---|---|---|---|---|---|---|---|---|
| | | Three doses, ≤ 60 days | | 6 | 101 | 0.12 | 0.04 | 0.31 |
| Vectored heterologous | Vectored | Two doses elegible | Infections | 2952 | 3531 | 1.00 | Ref | Ref |
| | | Two doses ineligible | | 1065 | 1203 | 1.01 | 0.90 | 1.12 |
| | | Three doses ≥60 days | | 455 | 660 | 1.21 | 1.05 | 1.40 |
| | | Three doses, ≤ 60 days | | 692 | 952 | 1.08 | 0.96 | 1.22 |
| | | Two doses elegible | Hospitalisations | 99 | 302 | 1.00 | Ref | Ref |
| | | Two doses ineligible | | 10 | 56 | 0.49 | 0.23 | 1.05 |
| | | Three doses ≥60 days | | 4 | 34 | 0.29 | 0.09 | 0.91 |
| | | Three doses, ≤ 60 days | | 7 | 65 | 0.28 | 0.12 | 0.65 |
| | | Two doses elegible | Death | 101 | 311 | 1.00 | Ref | Ref |
| | | Two doses ineligible | | 16 | 63 | 0.73 | 0.38 | 1.39 |
| | | Three doses ≥60 days | | 8 | 49 | 0.46 | 0.20 | 1.03 |
| | | Three doses, ≤ 60 days | | 6 | 95 | 0.18 | 0.08 | 0.45 |
| Vectored-mRNA | mRNA | Two doses elegible | Infections | 11790 | 13010 | 1.00 | Ref | Ref |
| | | Two doses ineligible | | 281 | 626 | 0.64 | 0.55 | 0.74 |
| | | Three doses ≥60 days | | 292 | 1233 | 0.90 | 0.75 | 1.09 |
| | | Three doses, ≤ 60 days | | 3016 | 7393 | 0.76 | 0.71 | 0.81 |
| | | Two doses elegible | Hospitalisations | 134 | 471 | 1.00 | Ref | Ref |
| | | Two doses ineligible | | 9 | 13 | 2.33 | 0.96 | 5.67 |
| | | Three doses ≥60 days | | 1 | 6 | 0.30 | 0.02 | 5.47 |
| | | Three doses, ≤ 60 days | | 7 | 140 | 0.09 | 0.04 | 0.24 |
| | | Two doses elegible | Death | 121 | 395 | 1.00 | Ref | Ref |
| | | Two doses ineligible | | 3 | 14 | 0.79 | 0.22 | 2.87 |
| | | Three doses ≥60 days | | 0 | 2 | | | |
| | | Three doses, ≤ 60 days | | 7 | 147 | 0.12 | 0.05 | 0.27 |
| Vectored-mRNA | Vectored | Two doses elegible | Infections | 11602 | 13091 | 1.00 | Ref | Ref |
| | | Two doses ineligible | | 263 | 631 | 0.61 | 0.53 | 0.71 |
| | | Three doses ≥60 days | | 1912 | 3009 | 1.09 | 1.02 | 1.17 |
| | | Three doses, ≤ 60 days | | 1856 | 2977 | 0.96 | 0.90 | 1.03 |
| | | Two doses elegible | Hospitalisations | 137 | 586 | 1.00 | Ref | Ref |
| | | Two doses ineligible | | 9 | 16 | 2.27 | 0.97 | 5.28 |
| | | Three doses ≥60 days | | 36 | 96 | 1.68 | 1.05 | 2.69 |

| | | | | | | | | |
|---|---|---|---|---|---|---|---|---|
| | | Three doses, ≤ 60 days | | 19 | 98 | 0.81 | 0.47 | 1.40 |
| | | Two doses elegible | Death | 121 | 484 | 1.00 | Ref | Ref |
| | | Two doses ineligible | | 3 | 20 | 0.49 | 0.13 | 1.78 |
| | | Three doses ≥60 days | | 18 | 78 | 0.94 | 0.52 | 1.69 |
| | | Three doses, ≤ 60 days | | 20 | 97 | 0.76 | 0.43 | 1.32 |

* Matching process used the nearest neighbour (1nn) matching, with up to five controls per case, based on age, gender, number of positive tests in the past, site of residence, presence or absence of comorbidities and number of previous tests.
† NA: Some odds ratios are not shown because of insufficient numbers of events
